# Predicting Alzheimer’s Trajectory: A Multi-PRS Machine Learning Approach for Early Diagnosis and Progression Forecasting

**DOI:** 10.1101/2023.11.28.23299110

**Authors:** Mashiat Mustaq, Naeem Ahmed, Sazan Mahbub, Clara Li, Yuichiro Miyaoka, TCW Julia, Shea Andrews, Md. Shamsuzzoha Bayzid, the Alzheimer’s Disease Neuroimaging Initiative

## Abstract

**INTRODUCTION:** Predicting the early onset of dementia due to Alzheimer’s Disease (AD) has major implications for timely clinical management and outcomes. Current diagnostic methods, reliant on invasive and costly procedures, underscore the need for scalable and innovative approaches. To date, considerable effort has been dedicated to developing machine learning (ML) based approaches using different combinations of medical, demographic, cognitive, and clinical data, achieving varying levels of accuracy. However, they often lack the scalability required for large-scale screening and fail to identify underlying risk factors for AD progression. Polygenic risk scores (PRS) have shown promise in predicting disease risk from genetic data. Here, we aim to leverage ML techniques to develop a multi-PRS model that captures both genetic and non-genetic risk factors to diagnose and predict the progression of AD in different stages in older adults.

**METHODS:** We trained and tested ML-based multi-PRS models, integrating genetically predicted clinical, behavioral, psychiatric, and lifestyle risk factors to predict the diagnosis of AD as well as the progression between different cognitive stages. We developed an automatic feature selection pipeline that identifies the relevant traits that predict AD. We also analyzed the interpretability of our proposed ML models and the selected features. Leveraging data from the Alzheimer’s Disease Neuroimaging Initiative (ADNI), Religious Orders Study and Memory and Aging Project (ROSMAP), and the IEU OpenGWAS Project, our study presents the first known end-to-end ML-based multi-PRS model for AD.

**RESULTS:** Relevant features were selected from an initial set of 53 polygenic risk scores computed for 1567 patients in the ADNI and 1642 patients in the ROSMAP dataset. The proposed multi-PRS ML method produced AUROC scores of 77% on ADNI and 72% on ROSMAP for predicting the diagnosis of AD, substantially surpassing the performance of the uni-variate PRS models. Our models also showed promise in predicting transitions between various cognitive stages (65%-75% AUROC scores). Moreover, the features identified by our automated feature selection pipeline are closely aligned with the widely recognized potentially modifiable risk factors for AD.

**DISCUSSION:** Multi-PRS-based machine learning models can identify risk factors and construct predictive models for early Alzheimer’s disease (AD) diagnosis. This approach offers an automated mechanism to harness genetic data for AD diagnosis and prognosis, enhancing our understanding of the role of various traits in AD development and progression. It will facilitate the implementation of preventive measures at an early stage, thereby contributing to more effective interventions and improved patient outcomes.

## 1 Introduction

AD presents a growing healthcare challenge, with its prevalence expected to rise globally [1]. Despite advancements in treatments, a cure remains elusive, necessitating further research into the disease’s etiology and risk factors [2]. AD manifests with diverse symptoms and trajectories, making accurate characterization crucial for effective treatments. Therefore, predicting AD is vital but hindered by the heterogeneity in patient populations and comorbidities.

There is no single test for diagnosing AD. Current medical tests involve the use of diagnostic tools in combination with medical history and other information, including neurological exams, cognitive testing, functional assessment, behavioral/mood evaluation, neuroimaging, cerebrospinal fluid, and/or blood tests to diagnose AD. However, these methods are time-consuming, costly, and not readily available, prompting the exploration of scalable alternatives. Machine learning (ML) algorithms have been developed [3–5], utilizing diverse data sets (e.g., including neuroimaging and CSF biomarkers, genotyping, demographic and clinical information, and cognitive performance), yet lack scalability and fail to capture underlying risk factors. Therefore, finding easily available and inexpensive markers capable of predicting the development and progression of AD in older adults has significant impacts on research and clinical care in the field.

Genetic methods offer robust alternatives to traditional biomarker-based disease predictions. Machine learning models incorporating Single Nucleotide Polymorphisms (SNPs) for AD have shown promise in clinical diagnosis and early detection [6, 7]. Genome-wide association studies (GWAS) identify genomic variants linked to diseases or traits by comparing genomes of affected and unaffected individuals. The increasing availability of genomic data and evidence of disease-associated variants fuels the popularity of GWAS. Polygenic Risk Scores (PRS) aggregate the effects of trait-associated variants discovered through GWAS, providing a nuanced understanding of disease risk beyond individual factors. By combining PRS with other influential attributes, a more accurate assessment of disease susceptibility is possible. Combining PRS with other attributes that influence disease risk can provide a more accurate picture of disease susceptibility than considering either factor alone.

While prior studies focused on uni-variate PRS models [8], integrating multiple PRS based on diverse traits enables comprehensive analysis, capturing the varied outcomes in Mild Cognitive Impairment (MCI, known as a transitional stage between normal aging and AD [9]), and thus allows improved AD progression prediction in older adults [10–14]. While some studies have utilized multi-PRS models to predict traits related to major psychiatric disorders and other conditions [15, 16], there is a lack of research exploring their application in predicting AD diagnosis and progression. The findings of Livingston et al. [17], indicating that approximately 40% of AD cases are attributable to 12 modifiable risk factors, are significant for understanding the preventable aspects of AD. Clinical risk scores (likelihood of developing a certain disease), which can be computed based on modifiable risk factors [18, 19], are essential for early identification of individuals at high risk and timely targeted interventions. However, not all datasets might contain the necessary variables for constructing these clinical risk scores. By integrating genetic data from various sources, multi-PRS-based approaches can potentially compensate for the lack of specific clinical variables/tests. Additionally, this approach enhances our existing knowledge about the genetic predispositions of AD, particularly the role of various risk traits in the development and progression of the disease.

Here, we present ML-based multi-PRS models, integrating genetically predicted clinical, lifestyle, and non-genetic risk factors to predict the diagnosis of AD as well as progression between different cognitive stages (i.e., Normal to MCI/AD and MCI to AD). We also investigated the efficacy of genetic data alone to diagnose and predict the progression of AD using ML-based techniques. Leveraging data from the ADNI, ROSMAP, and IEU OpenGWAS Project [20], our research presents a comprehensive approach. We employ an automatic feature selection pipeline, identifying relevant traits predictive of AD, and investigate the efficacy of genetic data in diagnosing and predicting AD progression. Our proposed methods are publicly available as an open-source project at https://github.com/Mashiatmm/AD Project Thesis.

## 2 Materials and Methods

### 2.1 Dataset

In this study, we used two datasets: ADNI [21] and ROSMAP [22] to build and assess our proposed multi-PRS ML models.

#### 2.1.1 ADNI

ADNI-1 is the initial study of ADNI while ADNI-GO, ADNI-2, and ADNI-3 are further phases, which not only included participants from previous phases for continued monitoring but also enrolled new participants for further investigation. SNP arrays of participants from different phases were combined to provide the data for this study. We refer to this combined dataset as the ADNI dataset. Overall, participants underwent medical, clinical, cognitive, functional, and behavioral assessments as well as neuroimaging and blood tests. Following the study visit, a study physician, who reviews the results of the assessments and other laboratory tests, will determine the best diagnosis (i.e., normal control, MCI, AD, or other) using criterion developed in the ADNI clinical protocol [23].

#### 2.1.2 ROSMAP

The ROSMAP dataset, which comprises data from 1,709 individuals, was generated in 2009 [24]. This dataset (SNP arrays) also includes samples from individuals, similar to the ADNI dataset. The diagnosis of AD in the ROSMAP dataset consists of a three-stage process including scoring of cognitive tests, clinical judgment by neuropsychologists, and diagnostic classification by clinicians [25, 26].

In our analysis, we focused on samples from both the ADNI and ROSMAP datasets with longitudinal data of clinical visits (a visit at the interval of every 6 months), enabling diagnoses to be made at multiple time points. We refer to positive samples as patients who were diagnosed with AD, while negative samples include patients with longitudinal data who did not have a diagnosis of AD within their medical records. The patients who at any stage of their follow-ups have been diagnosed with AD are the positive samples. However, when dealing with negative samples, there’s a possibility that individuals with limited longitudinal data might still develop Alzheimer’s later in life. For instance, a patient with only one year of longitudinal data (two visits) may develop Alzheimer’s within the following five years, even though this information is not present in our dataset. To address this issue, in addition to considering the entire dataset, we created subsets of negative samples where we excluded all negative samples with less than x years of available longitudinal data (where x = 2, 4, 6, 8). The purpose was to determine the most reliable longitudinal range for identifying negative cases. Table 1 displays the statistics of sample sizes associated with varying amounts of longitudinal data, following quality control and addressing population stratification, as elaborated in upcoming sections.

**Table 1:** Number of available samples in ADNI and ROSMAP dataset with different amounts of longitudinal data after quality control and population stratification

| Longitudinal data available | ADNI |  |  | ROSMAP |  |  |
| --- | --- | --- | --- | --- | --- | --- |
|  | Positive | Negative | Total | Positive | Negative | Total |
| All | 576 | 991 | 1567 | 655 | 987 | 1642 |
| 2 years | 576 | 729 | 1305 | 655 | 858 | 1513 |
| 4 years | 576 | 377 | 953 | 655 | 739 | 1394 |
| 6 years | 576 | 232 | 808 | 655 | 631 | 1286 |
| 8 years | 576 | 111 | 687 | 655 | 537 | 1192 |

### 2.2 Overview of the proposed method

Figure 1 shows the overall pipeline of our proposed multi-PRS-based model. Our primary dataset (to train and test our model) contains the SNP arrays from individuals within the ADNI and ROSMAP datasets. Quality control (QC) and population stratification (PS) techniques are employed to ensure the reliability of these datasets. Principal Components (PCs) were also generated from these datasets that will be utilized as covariates in subsequent analyses. Also, relevant traits, associated with AD, are identified using the existing literature and input from clinical experts. Next, GWAS data corresponding to these traits are obtained.

**Figure 1:**
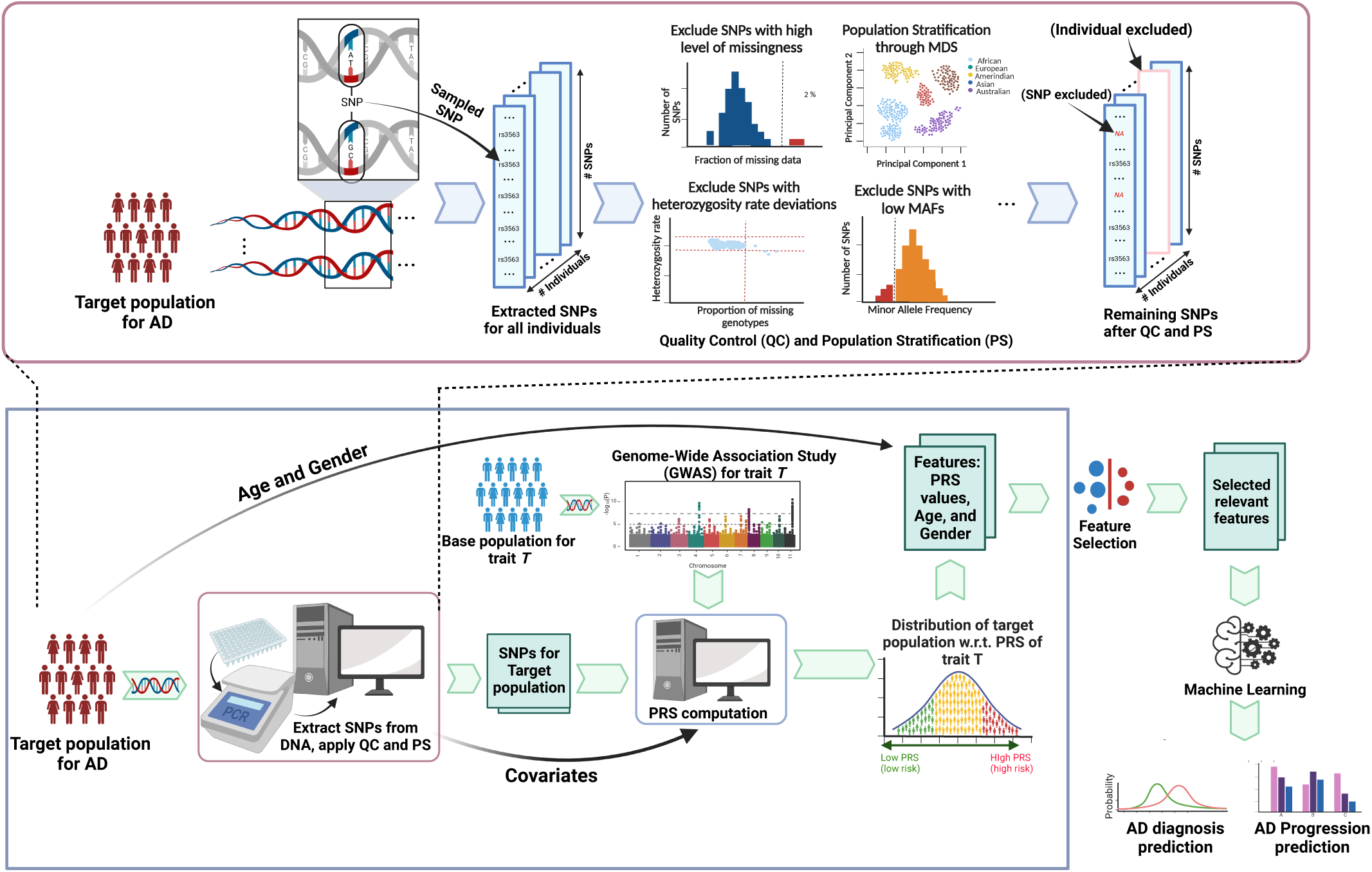
Schematic diagram of the multi-PRS-based pipeline for AD diagnosis and progression prediction. For a particular trait, genetic data of the base population and the target population go through the quality control and population stratification stages. Next, PRS are computed for the target population using the GWAS of the base population and genetic data (e.g. SNPs) of the target population. Next, the feature selection process is applied to the PRS of various traits along with two non-genetic features (age and gender). Finally, machine learning models are trained using the selected set of features.

PRS for a patient corresponding to a particular trait is computed using PRSice-2 [27], where we utilize the SNP arrays as target data and the GWAS data for that trait as the base data. We initially computed the PRS for 53 different traits which we use as features for our prediction model (the feature list is provided in Table S2 of the Supplementary Material). Additionally, we incorporate two non-genetic features, namely age, and gender. The obtained PRS for 53 traits, age, and gender - a total of 55 features are passed through a feature selection step, resulting in a much smaller set of traits. Subsequently, these selected features are utilized as features in our proposed ML-based prediction models. Quality Control, PRS generation, and feature selection steps are discussed in the subsequent sections.

### 2.3 Quality Control and Population Stratification

PRS of different traits for the target population are computed using the corresponding GWAS summary data. We leveraged the GWAS datasets generated by the IEU OpenGWAS Project [20]. We selected 53 traits based on expert opinions and existing literature [28–35], which were deemed significant in the context of AD development. The list of 53 traits is provided in the Supplementary Material (Table S2) which includes clinical traits (BMI, cholesterol, stroke, blood pressure, hypertension, hypothyroidism, etc.), behavioral traits (prospective memory), psychiatric traits (mental health issues, ADHD, loneliness, etc.), lifestyle traits (smoking tendency, food habits, sleep duration, etc). Among them, multiplem traits can be attributed to the 12 widely recognized modifiable risk factors reported in Livingstone et al. [17], such as less education, hypertension, air pollution, brain injury, smoking, physical inactivity, etc.

We utilized the Single Nucleotide Polymorphism (SNP) arrays of the individuals in the ADNI and ROSMAP datasets. SNP refers to a DNA sequence variation caused by the alteration of a single nucleotide (A/T/C/G) in the genome sequence. Before PRS generation, we conducted QC and PS steps of the SNP arrays using established guidelines [36,37] and the PLINK software [38]. QC checks included assessments for missingness of SNPs and individuals, sex discrepancies, Minor Allele Frequency (MAF) threshold deviations, deviation from Hardy-Weinberg Equilibrium (HWE), heterozygosity rate deviations, and relatedness of individuals. We performed exclusions based on several criteria: SNPs and individuals with substantial missingness (*>* 2%), cases of sex discrepancies, SNPs with low Minor Allele Frequency (MAF *<* 5%), and those not adhering to Hardy-Weinberg Equilibrium (HWE p-value *<* 10^−^6). Additionally, outliers in heterozygosity rate, deviating more than 3 standard deviations from the mean, were excluded. Individuals with high relatedness (pihat *>* 0.2) were addressed by removing the individual with the lower call rate.

For PS, we utilized the 1000 Genomes Project dataset [39] as a reference panel in the imputation process. We merged our target dataset (ADNI /ROSMAP) with the 1000 Genomes Project (1000GP) dataset and performed Multidimensional Scaling (MDS) to identify ethnic outliers. MDS analysis provided us with quantitative genetic components for each individual. To spot ethnic outliers, we compared our target samples’ component scores with those of the 1000 Genomes Project dataset, which we knew had ancestry information. By plotting these component scores alongside ancestry information, we observed that the majority of our target samples clustered with European ancestry. Samples outside this cluster, such as those from African and Asian ancestries, were identified as ethnic outliers, comprising less than 1% of the entire sample. They were excluded to prevent potential systematic bias. Consequently, our target dataset exclusively consisted of samples of European ancestry (for both ADNI and ROSMAP). We also generated ten covariates based on the MDS analysis, which were used as principal components in our PRS analysis. Thus, we obtained the quality controlled and population stratified genomic datasets which were used as input to PRSice-2 [27].

### 2.4 PRS Generation

The PRSice-2 software utilized the GWAS datasets as base data to extract summary statistics regarding the effective allele for the base phenotype. Then it identified the frequency of the effective allele within the target ADNI or ROSMAP data. By incorporating covariates and performing specific calculations [40], PRSice-2 generated Polygenic Risk Scores (PRS) for the base phenotype within the target dataset. The base GWAS dataset provides p-values for base SNPs represented as “LP” on the −*log*10 scale. To obtain the actual p-value for each base SNP, we used the formula: p val = 10^(−1∗^*^LP^* ^)^. These derived p-values served as the significance thresholds for the respective SNPs in our PRS calculation. For clumping, we used the default clumping parameter values in PRSice-2 (clump-kb = 250kb, clump-p = 1.00, and clump-r2 = 0.10).

### 2.5 Feature selection

Our initial feature set consists of PRS of 53 traits and two non-genetic features, age and gender. Here, age refers to the age at the beginning of the diagnosis of patients. To identify the most relevant traits for the prediction of AD, we employed automated feature selection processes using three different methods: filter, wrapper, and embedded methods [41, 42]. Filter methods, which are based on univariate statistical methods, were applied to the dataset using two techniques: information gain and correlation coefficient matrix. Additionally, we performed an assessment of the variance and mean absolute difference of each feature, with the intent of eliminating those that displayed a negligible variance or mean absolute difference. However, our analysis revealed no such features except *Age* which has a very low variance. Since most feature selection methods highlighted the importance of *Age*, and considering that the dataset mainly consisted of elderly probable AD patients, we did not exclude *Age* from our analysis.

The wrapper-based approach was utilized to optimize the subset of features for classification performance. This was achieved through the implementation of three techniques: Forward Feature Selection, Backward Feature Elimination [43], and Recursive Feature Elimination [44]. The embedded method combines the strengths of both filter and wrapper methods by considering the interaction of features in model training. In this method, the techniques employed were Lasso Regularization (L1) [45] and Random Forest Importance [46].

Each method employed in our selection process provided either a ranked position, or a numerical score for each feature, or in some instances, selected a subset of features. To ensure comparability, the ranks and scores were normalized and added as points corresponding to each feature. For methods that produced subsets of features, one point was added to each selected feature within the subset. Consequently, the cumulative points for each feature were computed and arranged in descending order of significance.

The XGBoost model was run on different top subsets of n features (e.g., top n features, where n = 1, 2, 3,…, n.) in the sorted list of features to identify the size of the most relevant subset. XGBoost was chosen since it showed promising results in previous works of multi-PRS prediction for different traits [16]. The subset of features with the best results was used for further experiments.

### 2.6 ML models and evaluation metrics

We employed four different types of ML models, Support Vector Machine (SVM), Random Forest (RF), Extreme Gradient Boosting (XGBoost or XGB), and Neural Network (NN) to predict AD from our datasets. We used 10-fold cross-validation to ensure the reliability of our models’ performance. The NN consists of four hidden layers and thirty-two nodes. We could not effectively use any deep learning models due to the small size of the dataset [47]. The proposed models were assessed based on a wide array of evaluation metrics such as precision, recall, F1-score, area under the receiver operating characteristic curve (AUROC), and area under the precision-recall curve (AUPRC). We fixed the random seed to 42 for reproducibility.

## 3 Results

### 3.1 Diagnosis of AD using a multi-PRS-based ML model

#### 3.1.1 Longitudinal data analysis

As discussed in Sec. 2.1, we considered different model conditions with different amounts of longitudinal data to assess how variations in the availability of longitudinal data affect the performance of our predictive model. It is expected that ML models will perform better with increasing numbers of data samples. Likewise, having longer amounts of longitudinal data for each patient will help the ML models learn and predict the diagnosis/progression of AD. However, there is a trade-off between these two – as we increase the threshold of the longitudinal range, the number of samples that meet the criteria for the extended timeframe gradually reduces (for both ADNI and ROSMAP). We note that the number of AD-positive samples in different longitudinal ranges remains the same for both ADNI and ROSMAP datasets. However, the number of negative samples decreases with increasing longitudinal ranges To remove the biasness (towards positive or negative samples) in ML models, it is required to use a balanced dataset by taking an equal number of positive and negative samples. But it results in the reduction of the total dataset size. In the case of ADNI, when we consider patients with a minimum longitudinal data of 2 years or less, the number of negative samples exceeds that of positive samples. The same trend is observed in the ROSMAP dataset when the minimum threshold for longitudinal data is 4 years or less. Consequently, in these specific ranges for each dataset, when balancing by equalizing the number of negative and positive samples, the dataset size remains the same, and the model’s performance remains consistent. For other cases, the balanced dataset size decreases with the increasing longitudinal range. Our target is to find as long a threshold as possible that will result in a reasonably large number of data samples. To accomplish this, we analyzed different model conditions with varying longitudinal ranges using the XGBoost algorithm.

The results shown in Figure 2 indicate that the performance of ML models improves with an increasing longitudinal data range, despite the decreasing number of patients within a particular range. This observation suggests that longitudinal data with a relatively long time frame substantially reduces the chances of misclassifying a potential positive sample as negative. Nonetheless, as shown in Figure 2, when the longitudinal range reached 8 years, the number of available samples in the ADNI dataset after making it balanced dropped down to *<* 250. So we set the longitudinal range of 6 years for the ADNI dataset. In contrast, the ROSMAP dataset maintained a sufficient number of balanced samples even when considering an 8-year longitudinal range. Therefore, we used an 8-year longitudinal range for the ROSMAP dataset.

**Figure 2:**
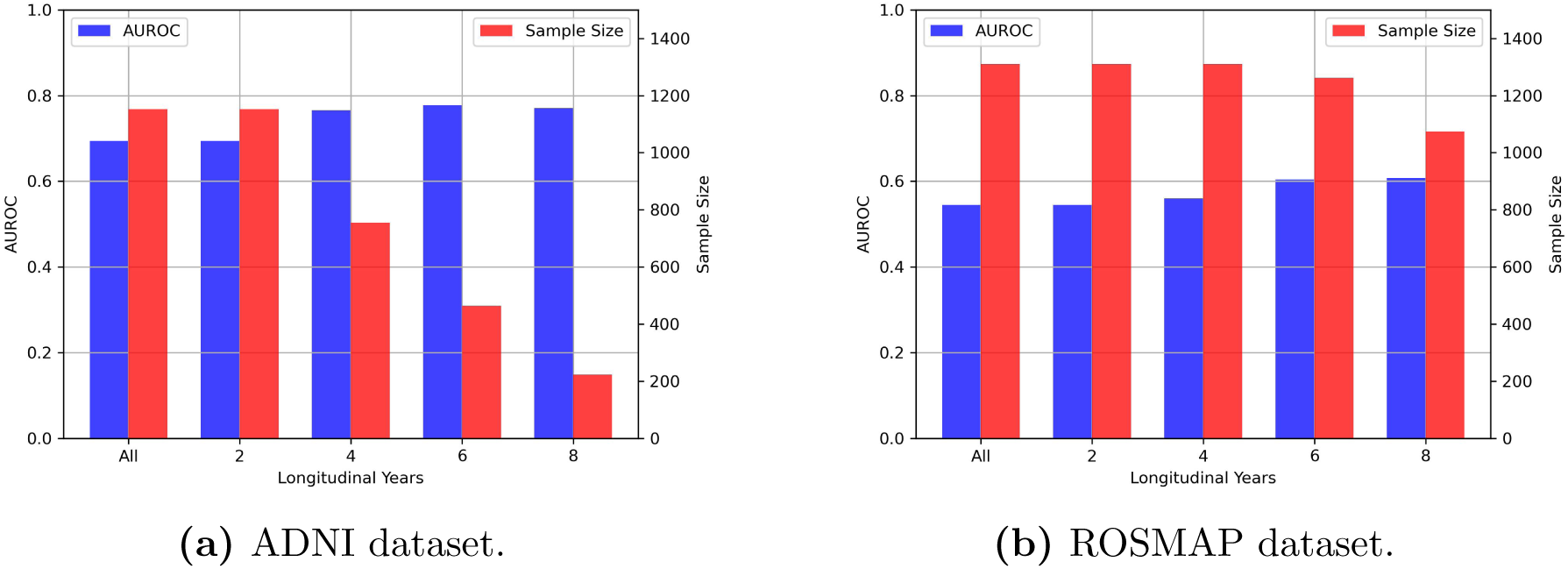
Analysis of different ranges of longitudinal data in ADNI and ROSMAP datasets. For different ranges (e.g, All-data, 2, 4, 6, and 8 years), we show the number of patients within those ranges and the performance of the XGBoost predictor in terms of AUROC scores.

#### 3.1.2 Feature selection

Next, we tried to find an optimal set of PRS features that are most predictive of AD. The feature selection pipeline (described in Sec. 2.5) was executed for both datasets, generating rankings of the initial pool of 55 features based on their relevance/impact on AD diagnosis. The ordered features with their cumulative points are shown in Figures 3 and 4 for ADNI and ROSMAP datasets, respectively. Remarkably, the outcomes of our automatic feature selection pipeline are aligned with the 12 widely recognized modifiable risks for AD [17] (provided in Table S3 in the supplementary materials). Specifically, our pipeline placed various features, relevant to these 12 factors, such as hypertension, hearing difficulty, physical inactivity, low social contact, alcohol consumption, and smoking in the top 20 features for ADNI and ROSMAP datasets, respectively. Notably, even the risk factors that did not make it into the top 20 features (such as obesity, diabetes, brain injury, and air pollution) exhibited significantly high cumulative feature selection scores, indicating the effectiveness of our proposed selection process. We note that features related to less education and depression received low feature selection scores for ADNI and ROSMAP, respectively, even though these two traits are among the 12 risk factors.

**Figure 3:**
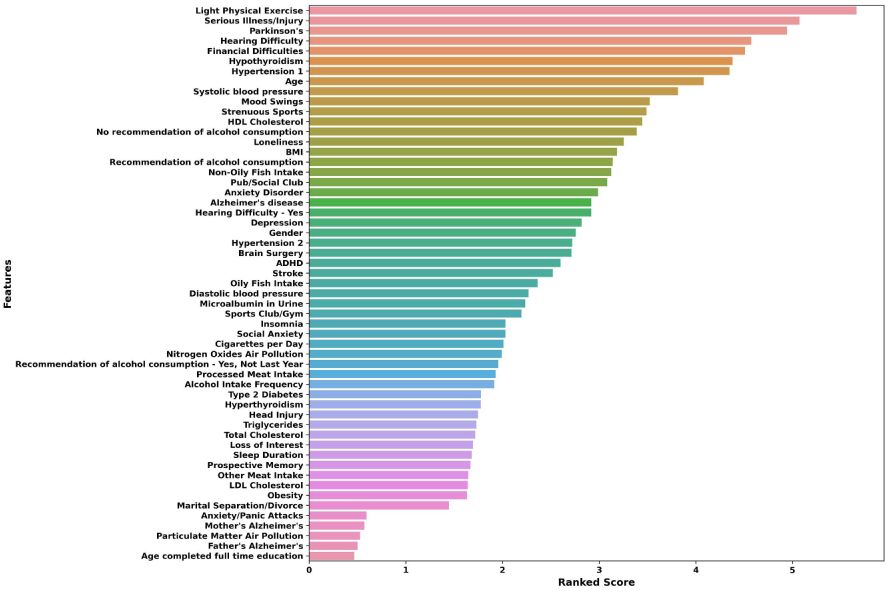
Sorting the set of features based on their cumulative points obtained from the feature selection process for the ADNI dataset.

**Figure 4:**
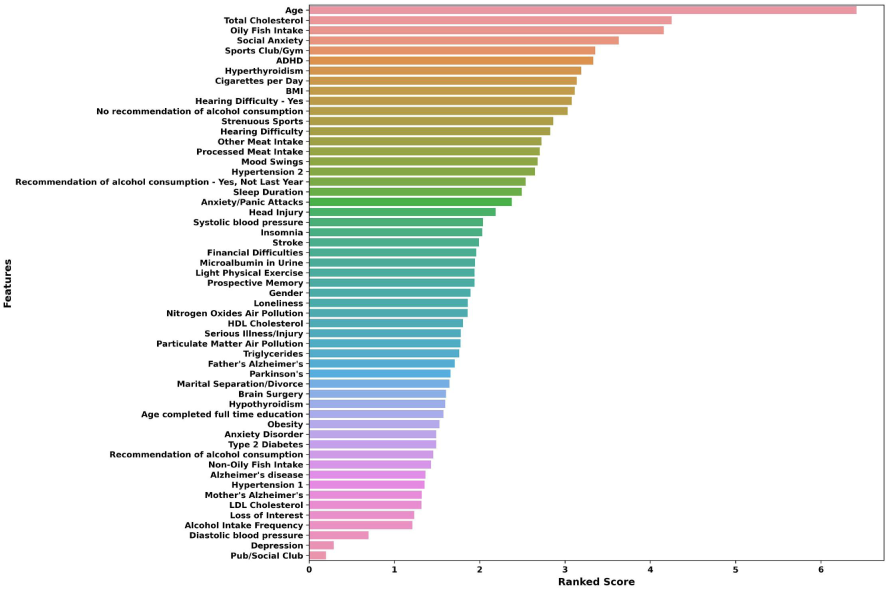
Sorting the set of features based on their cumulative points obtained from the feature selection process for the ROSMAP dataset.

Next, we investigated the performance of an ML prediction model on different subsets of features with varying sizes, where the features are selected one by one based on their priorities. For instance, when considering a subset size of 10, we selected the top 10 features according to the ranking produced by our feature selection pipeline. In Figure 5, we show the AUROC scores for different subset sizes.

**Figure 5:**
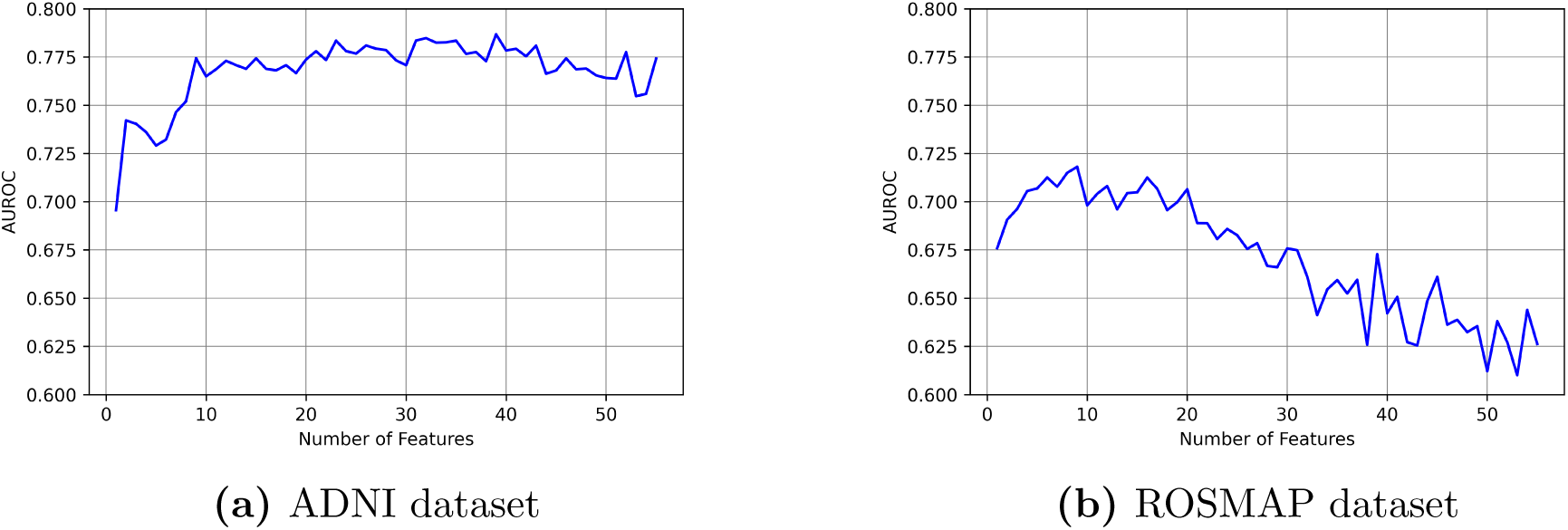
Finding optimal sets of features from an initial set of 55 features. We investigated the AUROC values for different subsets of features. For a subset of n features, we choose the top n feature from the ranked list of features generated by our feature selection process.

For the ADNI dataset, the AUROC score increased rapidly with increasing feature set size and reached 0.77 with 9 features (Figure 5a). However, as we further increased the number of features, the AUROC score did not significantly improve but rather stabilized within the range of 0.75 to 0.78, reaching its peak at 0.78 when employing 39 features. Conversely, on the ROSMAP dataset, the AUROC initially improved as the number of features increased, reaching its peak at 0.72 with 9 features. However, beyond this point, the AUROC started to decline (Figure 5b). These different trends observed on the two datasets could be attributed to the different impacts of various traits of ADNI and ROSMAP on AD prediction. Figures 3 and 4 indicate that for the ADNI dataset, many features showed almost similar significance to the prediction of AD and the cumulative feature selection points dropped relatively slowly in the sorted list. This could partly contribute to the stabilization of the AUROC beyond 9 features on the ADNI dataset. In contrast, for the ROSMAP dataset, the significance of features substantially decreased beyond the initial few features. Consequently, expanding the feature set involved incorporating poorly significant features into the prediction model, leading to a decline in the overall performance beyond the top 9 features. Hence, we used 9 top features for both ADNI and ROSMAP datasets from the ordered lists shown in Figures 3 and 4, respectively (these lists are provided in Supplementary Material: Table S4).

#### 3.1.3 Performances of different ML models in AD diagnosis

Table 2 presents a comparison of four multi-PRS-based ML models in predicting AD on the ADNI, ROSMAP, and a dataset with both ADNI and ROSMAP. The reported performance metrics are based on samples with a minimum of six years of longitudinal data for ADNI and eight years for ROSMAP. For the merged dataset of ADNI and ROSMAP (referred to as the combined dataset), samples with at least six years of longitudinal data for both were combined. Since the number of negative samples was relatively low, an equal number of positive samples was randomly selected to ensure a balanced dataset before the training.

**Table 2:** Comparison of performances of different ML models on ADNI, ROSMAP, and the combined datasets.

| Dataset | Model | Accuracy | Precision | Recall | AUROC | AUPRC | F1-score |
| --- | --- | --- | --- | --- | --- | --- | --- |
| ADNI | NN | 0.777 | 0.825 | 0.576 | 0.762 | 0.649 | 0.776 |
|  | SVM | 0.727 | 0.729 | 0.720 | 0.765 | 0.724 | 0.756 |
|  | RF | 0.736 | 0.757 | 0.698 | 0.767 | 0.733 | 0.777 |
|  | XGB | 0.728 | 0.746 | 0.693 | 0.774 | 0.725 | 0.794 |
| ROSMAP | NN | 0.619 | 0.430 | 0.434 | 0.547 | 0.408 | 0.557 |
|  | SVM | 0.653 | 0.657 | 0.656 | 0.729 | 0.651 | 0.745 |
|  | RF | 0.661 | 0.678 | 0.623 | 0.709 | 0.659 | 0.711 |
|  | XGB | 0.658 | 0.676 | 0.616 | 0.718 | 0.656 | 0.725 |
| Combined | NN | 0.610 | 0.616 | 0.426 | 0.587 | 0.481 | 0.574 |
|  | SVM | 0.642 | 0.639 | 0.662 | 0.696 | 0.649 | 0.676 |
|  | RF | 0.630 | 0.633 | 0.627 | 0.689 | 0.628 | 0.669 |
|  | XGB | 0.638 | 0.635 | 0.655 | 0.690 | 0.643 | 0.675 |

We achieved an AUROC score as high as 77% for the ADNI dataset and 72% for the ROSMAP dataset. For the combined dataset, our feature selection pipeline selected 10 features, which yielded an AUROC score of around 69%. The F1-scores obtained also fell within a similar range. We note that multi-PRS-based models performed better on ADNI than ROSMAP and the combined dataset, despite ADNI having lower numbers of samples and being more imbalanced than ROSMAP. This variation can be due to the varying significance of the features concerning the diagnosis of AD. As illustrated in Figures 3 and 4, the initial features exhibited higher cumulative points in the ADNI dataset in contrast to the ROSMAP dataset. For ROSMAP, the top feature was *Age* and the cumulative feature selection points declined considerably for most features afterward. Consequently, the diagnosis of AD in the ROSMAP dataset may not be as closely correlated with the available features as it is in the ADNI dataset. Similarly, combining datasets (thereby increasing the number of samples) did not result in improved performances. This could be due to the demographic and other differences between the ADNI and ROSMAP datasets (see Section 2 of Supplementary Material for details).

As shown in Table 2, the performance of the NN model differs depending on the dataset it is applied to. When we applied it to the ADNI dataset, it produced better results in terms of accuracy only. However, when we applied it to the ROSMAP and Combined datasets, the scores dropped significantly. In contrast, other models like SVM, RF, and XGB showed consistent performance across all datasets. XGBoost performed as well as or better than other models across all these three datasets. Therefore, we used the XGBoost model for our further analyses.

#### 3.1.4 The impact of the AD PRS in predicting the risk of AD

PRS for AD (AD-PRS) is calculated using SNPs with Alzheimer’s disease association. Due to the polygenicity of AD, AD-PRS has been shown to be successful in AD risk prediction [11, 48, 49]. However, our feature selection process did not identify AD-PRS (out of the initial pool of 53 PRS features) as an important feature so we did not include AD-PRS in our ML models. In this section, we analyzed the effect of including AD-PRS by running our ML models with and without AD-PRS. The results are presented in Figure 6.

**Figure 6:**
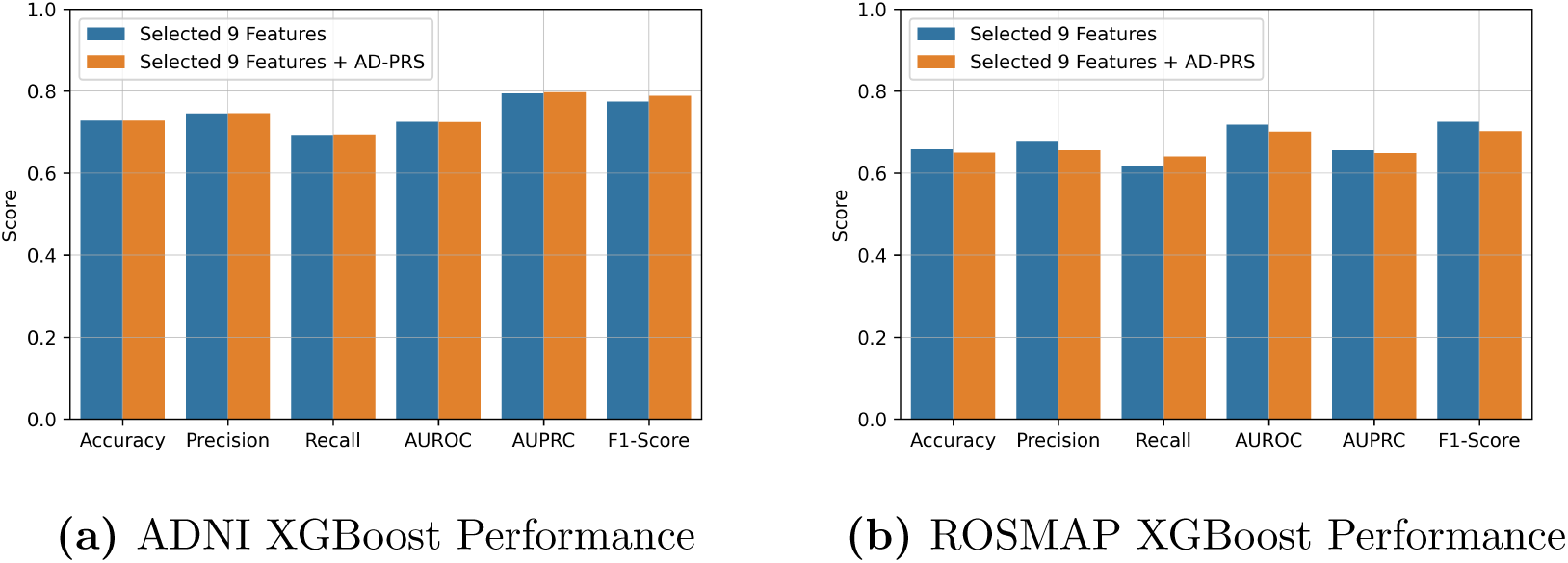
Performance comparison of XGBoost with and without the AD-PRS feature.

The results show that the inclusion of the AD-PRS does not make any notable changes in the performance of our multi-PRS-based ML models on these particular datasets. To further investigate this, we assessed the power of the AD-PRS alone in diagnosing AD. In this regard, we evaluated the efficacy of our multi-PRS model in contrast to the single PRS (AD-PRS) model. We implemented the following two AD-PRS-based approaches:

1. **Thresholding based on AD-PRS:** In this method, we considered the AD-PRS values of the samples within our balanced dataset (6 years for ADNI and 8 years for ROSMAP). Subsequently, we computed the mean of the PRS values of the positive samples and employed it as a threshold to classify samples as positive or negative. Samples with PRS values exceeding the threshold were predicted as positive, while those below were classified as negative.
2. **AD-PRS based ML model:** an ML model with only AD-PRS as a genetic feature along with sex and gender as non-genetic features (similar to prior studies [11, 48, 49]).

In Table 3, we show a comparison between our multi-PRS models with these single PRS strategies. These results show that the multi-PRS-based models perform substantially better than single-PRS-based models, except for the ROSMAP dataset where the single-PRS-based ML model achieves a comparable AUROC score as the multi-PRS model. Unlike previous studies, which showed remarkable accuracy for single-PRS-based (i.e., AD-PRS) ML models on the IGAP dataset [49], we found that AD-PRS alone performs poorly on the ADNI dataset. In particular, the threshold-based approach did not show notable predictive power. Thus, the performance of single-PRS-based (and multi-PRS-based) models may substantially vary across different datasets. Therefore, selecting suitable methods becomes particularly challenging when the data are heterogeneous, which is often so for medical data.

**Table 3:** Performance of multi-PRS and single-PRS-based models. We show the AUROC values of different approaches. We used the XGBoost method as the predictive model.

| Dataset | Experiment | AUROC |
| --- | --- | --- |
| ADNI | Selected 9 Features | 0.774 |
|  | Thresholding AD-PRS | 0.599 |
|  | AD-PRS + Age + Gender | 0.576 |
| ROSMAP | Selected 9 Features | 0.718 |
|  | Thresholding AD-PRS | 0.509 |
|  | AD-PRS + Age + Gender | 0.690 |

#### 3.1.5 Predicting AD with genetic features alone

To investigate the predictive power of genetic features alone, we omitted age and gender from the feature set and conducted our feature selection process using only the 53 PRS. We then trained the XGBoost model, incrementally introducing genetic features in order of their significance. In Figure 7, we present a visual comparison of the model’s performance when including and excluding age and gender as features. It is worth noting that the subsets of features displayed on the x-axis are distinct for the cases with and without age and gender.

**Figure 7:**
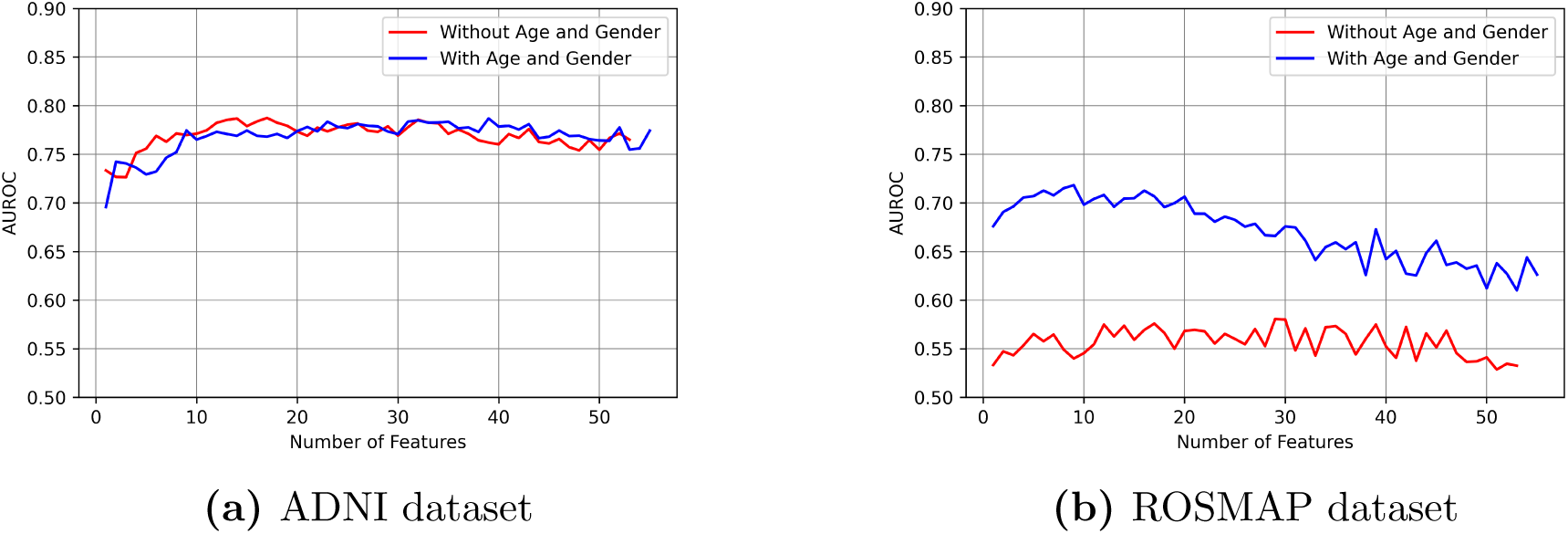
Performance of the predictive models with and without the non-genetic features age and gender. We show the AUROC values for different subsets of ranked features for both ADNI and ROSMAP datasets.

In the case of ADNI, the AUROC score consistently remained within the range of 77-78% when considering only genetic features. This suggests that, on this particular dataset, the genetic features alone produce results comparable to those obtained with the addition of non-genetic features. However, on the ROSMAP dataset, the AUROC score decreased significantly after excluding age and gender. This decline can be primarily attributed to the significant impact of age on the prediction of AD in ROSMAP, as evidenced by its high rank shown in Figure 4.

#### 3.1.6 Impact of APOE

We also examined the impact of including genetic features related to the APOE gene, which is known to increase the risk of developing AD [50]. However, we observed that the performance of the ML models did not improve with the addition of these features for both ADNI and ROSMAP. We present the result in Section 8.1 of Supplementary Materials.

#### 3.1.7 Interpretability of the proposed models

ML models often suffer from a lack of interpretability, making it hard to understand how they make predictions. Therefore, it is important to summarize the reasons for the network behavior, or produce insights about the causes of their decisions and thus gain the trust of users. Our feature selection pipeline, as discussed in Sec. 2.5, selected a suitable set of features to predict AD. To investigate how much each feature contributes to the model’s prediction, we applied the SHAP (SHapley Additive exPlanations) [51] framework to our trained XGBoost model. The framework generates SHAP values that measure the impact of variables, taking into account the interaction with other variables by comparing what a model predicts with and without the feature. SHAP values provide valuable insights into the significance of individual features and their effects on the model’s outcomes.

The SHAP values obtained on ADNI and ROSMAP datasets are shown in Figure 8. On the y-axis, features are arranged in order of importance from top to bottom, while the x-axis illustrates the corresponding SHAP values. Features transitioning from blue to red with the increasing SHAP values indicate a positive contribution towards the model’s output, indicating their role in pushing the output toward higher values. Conversely, features transitioning from red to blue demonstrate a negative correlation with the trained model. The magnitude of a SHAP value signifies the strength of the positive or negative effect.

**Figure 8:**
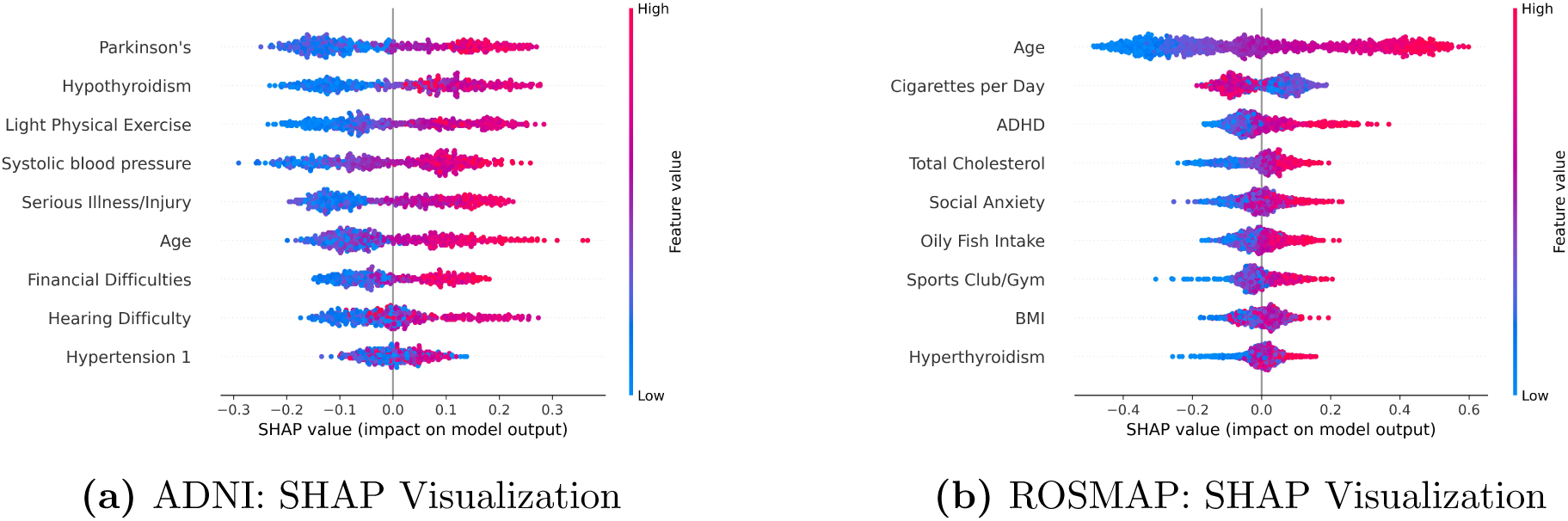
SHAP-based visualization of the impact of different features on the outcome of the XGBoost model for the ADNI and ROSMAP datasets.

The SHAP values indicate that each of the selected 9 features for the ADNI dataset positively affects the outcome of the XGBoost model, meaning that the higher values of these features contribute to the positive diagnosis of AD. However, the feature of *Light Physical Exercise* should have a negative impact on the diagnosis of AD, indicating that higher values of genetically predicted *Light Physical Exercise* should be associated with a reduced likelihood of an AD diagnosis. Therefore, the model’s behavior in terms of this particular feature is not interpretable. For the ROSMAP dataset, the magnitude of the SHAP values is less than those on the ADNI dataset. Moreover, *Cigarettes per Day* and *Sports Club/Gym* are not correctly correlated with the model’s outcome. For instance, according to the SHAP values, higher consumption of cigarettes/day tends to negatively impact the output (i.e., negative diagnosis of AD) – which should not be the case and thus is not interpretable. Similarly, higher sports club/gym tends to contribute to the positive diagnosis of AD – which should be the opposite. This explains the relatively low performance of the ML models on the ROSMAP dataset compared to the ADNI dataset. Another observation is that the order of significance for features for both datasets in terms of the SHAP values did not completely match the rank we obtained in our feature selection process.

### 3.2 Predicting progression towards AD using multi-PRS based ML models

In our next experiment, we investigated the power and efficacy of multi-PRS-based models in predicting progression cases from one cognitive stage to others. We specifically explored the following three cases.

1. Progression from non-AD states (CN or MCI) to AD
2. Progression from the CN state to either MCI or AD
3. Progression from the MCI state to AD

We first identified individuals based on their initial diagnosis, which corresponds to the diagnosis they received during their first visit. We then categorized the samples into two groups: positive and negative cases based on their final diagnosis. For instance, if a person was initially diagnosed as CN during the first visit and later received a final diagnosis of MCI or AD, they were considered a positive sample indicating disease progression. Conversely, if their final diagnosis did not indicate MCI or AD, they were considered a negative sample, signifying no progression from their initial state. We paid attention to the longitudinal range of the data to ensure the reliability of our predictions and excluded samples with less than 4 years of data in the ADNI dataset and less than 8 years of data in the ROSMAP dataset. Like our approach in predicting the final diagnosis (as in Sec. 3.1), we determined the optimal number of features to consider by investigating the AUROC scores vs. the number of features (provided in Section 6 of the Supplementary materials). For each of the three considered progression cases, we conducted training and testing on both the ADNI and ROSMAP datasets, with the number of features determined by our feature selection pipeline.

We evaluated the effectiveness of the multi-PRS-based approach in predicting AD progression using the XGBoost algorithm. Various performance metrics were measured, including accuracy, precision, recall, AUROC, AUPRC, and F1-score. The results, along with the sample size and the total number of considered features, are presented in Table 4.

**Table 4:** Table summarizing Performance Metrics for Alzheimer’s Disease Progression Prediction using the Multi-PRS-Based Approach with the XGBoost Algorithm. The table presents key evaluation metrics including accuracy, precision, recall, AUROC, AUPRC, and F1-score. It also provides information on sample size and the total number of features considered. The analysis focuses on progression cases and encompasses evaluations across both ADNI and ROSMAP datasets. The number of features is determined from the best AUROC results, and the dataset includes samples with 6 years of longitudinal data for ADNI and 8 years of longitudinal data for ROSMAP.

| Progression From | Dataset | Sample Size | Total Features | Accuracy | Precision | Recall | AUROC | AUPRC | F<sub>1</sub>-score |
| --- | --- | --- | --- | --- | --- | --- | --- | --- | --- |
| Non-AD (CN/MCI) | ADNI | 562 | 12 | 0.703 | 0.732 | 0.652 | 0.753 | 0.742 | 0.701 |
|  | ROSMAP | 1074 | 5 | 0.652 | 0.658 | 0.635 | 0.705 | 0.705 | 0.645 |
| CN | ADNI | 128 | 5 | 0.564 | 0.549 | 0.587 | 0.596 | 0.645 | 0.546 |
|  | ROSMAP | 730 | 7 | 0.663 | 0.667 | 0.657 | 0.698 | 0.671 | 0.659 |
| MCI | ADNI | 354 | 28 | 0.672 | 0.713 | 0.573 | 0.753 | 0.763 | 0.666 |
|  | ROSMAP | 696 | 15 | 0.617 | 0.628 | 0.604 | 0.682 | 0.677 | 0.606 |

Considering the inherent difficulty of predicting progression between cognitive stages and that we focused mostly on genetic data with limited numbers of training samples with short longitudinal ranges of patient data, achieving AUROC and F1-scores in the range of 65% to 75% is remarkable. An exception is seen in the case of progression from the CN State evaluated on the ADNI data, where the sample size is considerably smaller, resulting in relatively poor and inconsistent performance scores. Also, though the number of features to obtain the best results reported in Table 4 for the progression from MCI cases to AD is relatively high, our feature selection process shows that the performance remains consistent after adding 10 features (see Section 6 of supplementary materials). Therefore, for the progression from MCI to AD, using at least ten features achieves similar performance scores.

## 4 Conclusions

In this study, we aimed to develop fully automated and highly accurate ML-based methods to predict AD and its progression, which is currently only partially addressed through ML models using various clinical and imaging data. Our methods leverage the power of multi-PRS-based models, utilizing readily accessible GWASs – thereby understanding the role that genetics plays in the onset and progression of AD.

Existing tests to diagnose and assess the progression of AD are mainly based on various expensive and/or invasive methods. In contrast, PRS summarizes the estimated effect of many genetic variants on an individual’s phenotype and thus provides further insight into AD risk and progression. In particular, data from thousands of GWAS are now available. However, the state-of-the-art in developing clinically translatable frameworks to assess one’s susceptibility to a particular disease by utilizing PRS leaves much to be desired. Our proposed method is the first of its kind to predict AD diagnosis and progression using a multi-PRS-based ML model. We aimed to effectively use genetic data to assess AD as well as to elucidate the role of various traits in the diagnosis and progression of the disease.

ML-based algorithms allow us to analyze a plethora of data and make a prediction based on different features, in addition to capturing interactions between various features, which is impossible manually otherwise. Moreover, these algorithms try to adaptively select the best set of parameters, thus removing human biases and the necessity of manual intervention while setting various parameters. These underscore the importance of developing ML-based models. Our experimental results on the ADNI and ROSMAP datasets indicate that the proposed multi-PRS-based ML models have merit in the evaluation of AD. We believe our proposed ML-based approach will evolve with the availability of new data (e.g., GWAS for new traits, new large-scale longitudinal datasets like ADNI and ROSMAP, etc.) and in response to scientific findings and medical experts’ feedbacks – laying a firm and broad foundation for fully automated, highly accurate and clinically translatable multi-PRS-based prediction models for AD diagnosis and progression. Our approach also can be extended to identify unique traits associated with AD diagnosis and progression in ethnic/racial minority groups if such a dataset becomes available in the future.

This study makes several notable contributions. This is the first known study to utilize multi-PRS-based ML models in AD diagnosis and progression prediction. Our results on the ADNI and ROSMAP datasets suggest that leveraging a multi-PRS model may capture the risk of AD better than the AD-PRS alone. However, this should be confirmed by assessing the performance of single- and multi-PRS models on more datasets, which we left as future work. We presented a rigorous feature selection process to select a suitable set of features that are most predictive of AD, thereby enhancing our understanding of the etiology of AD. We also assessed the impact of using only genetic features (without important non-genetic features such as age and gender). Our results suggest that, on some datasets, the genetic features alone may produce results comparable to those obtained with the addition of non-genetic features. Furthermore, we attempted to analyze the interpretability of the behavior of the ML models and our proposed feature selection process.

Developing ML models to predict transitions between cognitive stages is another notable contribution of this study. This study shows the power of multi-PRS-based models in predicting the progression toward AD from the CN and MCI states. However, these results also underscore the presence of ample room for enhancement, indicating that effectively predicting the transition toward AD using multi-PRS-based models leaves much to be desired. Since our approach demonstrated poor performance when the sample size was too small between two different states, the availability of a larger number of samples between these states could enhance its performance. Subsequently, we find that developing AD progression models, far from being a ‘solved problem’, would benefit from new attention and therefore should propel new future research directions. One potential approach could involve measuring the decline in standardized cognitive scales (such as ADAS, CDR etc) using regression models rather than focusing solely on the transition between two different states. Such a scale may offer a more accurate estimate of deterioration over time.

This study has some limitations and can be extended in several directions. Our analysis predominantly relied on the ADNI and ROSMAP datasets, which primarily consist of individuals of European ancestry with limited diversity. The population outliers of different races were dropped during the population stratification steps, which resulted in a dataset with samples from only the European ancestry. Hence, it is essential to compile and analyze additional datasets encompassing a more comprehensive range of ancestral backgrounds to ensure that the reported performance is not specific to the patient cohorts presented here. As such, the results presented here need to be interpreted with caution, recognizing that the generalization of these results may require more extensive cross-validation. Developing an unbiased multi-PRS model to predict AD across ancestral groups is important for the generalization of the results. Therefore, future studies need to develop cross-ancestry multi-PRS ML models by leveraging ancestry informative methods to predict AD in older adults across diverse ancestral groups.

The sample sizes in the ADNI and ROSMAP datasets are not large enough to leverage advanced deep learning techniques. Therefore, compiling much larger datasets and investigating the performance of various deep learning techniques will be an important research avenue. Our study achieved AUROC scores in the range of 60-80%, which we believe will be improved with larger datasets such as the International Genomics of Alzheimer’s Project (IGAP), which will enable training deeper networks. Apart from that, the SHAP analysis revealed that certain features had an impact on our model that contradicted the existing literature. Also, the performance on ROSMAP is not as good as that of ADNI, which we believe is due to the features having low correlation with the samples of ROSMAP. Also, the distinction of performance can be attributed to the different diagnostic procedures for both datasets. Therefore, there is a need for increased emphasis on correctly interpreting the significance of each feature in our trained model and improving its performance for various datasets. Additionally, there are emerging evidence suggesting that AD likely develops from several factors, including genetic, lifestyle, and environmental factors [52, 53]. However, this study is limited to genetic traits from a predominantly non-Hispanic White population. Future studies can aim to examine the interactions between the genetic and non-genetic risk factors across different racial/ethnic groups.

## Data Availability

Data used in the preparation of this article are available from the Alzheimer's Disease Neuroimaging Initiative (adni.loni.usc.edu) and Religious Orders Study and Memory and Aging Project (https://www.radc.rush.edu) databases.

## Notes

### Competing Interest Statement

The authors have declared no competing interest.

### Funding Statement

This study was supported by the Interstellar Early Career Investigator Award (jointly presented by the New York Academy of Sciences (NYAS) and the Japan Agency for Medical Research and Development (AMED)). It was also partially supported by the Research and Innovation Centre for Science and Engineering at BUET (RISE-BUET) Internal Research Grant (ID: 2021-01-016).
Data collection and sharing for this project was funded by the Alzheimer's Disease Neuroimaging Initiative (ADNI) (National Institutes of Health Grant U01 AG024904) and DOD ADNI (Department of Defense award number W81XWH-12-2-0012). ADNI is funded by the National Institute on Aging, the National Institute of Biomedical Imaging and Bioengineering, and through generous contributions from the following: AbbVie, Alzheimer's Association; Alzheimer's Drug Discovery Foundation; Araclon Biotech; BioClinica, Inc.; Biogen; Bristol-Myers Squibb Company; CereSpir, Inc.; Cogstate; Eisai Inc.; Elan Pharmaceuticals, Inc.; Eli Lilly and Company; EuroImmun; F. HoffmannLa Roche Ltd and its affiliated company Genentech, Inc.; Fujirebio; GE Healthcare; IXICO Ltd.; Janssen Alzheimer Immunotherapy Research & Development, LLC.; Johnson & Johnson Pharmaceutical Research & Development LLC.; Lumosity; Lundbeck; Merck & Co., Inc.; Meso Scale Diagnostics, LLC.; NeuroRx Research; Neurotrack Technologies; Novartis Pharmaceuticals Corporation; Pfizer Inc.; Piramal Imaging; Servier; Takeda Pharmaceutical Company; and Transition Therapeutics. The Canadian Institutes of Health Research is providing funds to support ADNI clinical sites in Canada. Private sector contributions are facilitated by the Foundation for the National Institutes of Health (www.fnih.org). The grantee organization is the Northern California Institute for Research and Education, and the study is coordinated by the Alzheimer's Therapeutic Research Institute at the University of Southern California. ADNI data are disseminated by the Laboratory for Neuro Imaging at the University of Southern California.
The ROSMAP studies were funded by the National Institute on Aging: P30AG010161 ADCC, P30AG072975 ADRC, R01AG015819 RISK, R01AG017917 MAP, U01AG46152 AMP-AD Pipeline I, U01AG61356 AMP-AD Pipeline II.

### Author Declarations

Data used in the preparation of this article are available from the Alzheimer's Disease Neuroimaging Initiative (adni.loni.usc.edu) and Religious Orders Study and Memory and Aging Project (https://www.radc.rush.edu) databases. Accessing the datasets required special access requests to be approved by the corresponding authority.

### Summary of Updates

An additional literature review has been conducted, resulting in an updated introduction. Furthermore, grammatical errors have been corrected and the text has been appropriately rephrased.

